# The effect of Alzheimer’s disease-associated genetic variants on longevity

**DOI:** 10.1101/2021.02.02.21250991

**Authors:** Niccolò Tesi, Marc Hulsman, Sven J. van der Lee, Iris E. Jansen, Najada Stringa, Natasja M. van Schoor, Philip Scheltens, Wiesje M. van der Flier, Martijn Huisman, Marcel J. T. Reinders, Henne Holstege

## Abstract

The genetics underlying human longevity is influenced by the genetic risk to develop -or escape- age-related diseases. As Alzheimer’s disease (AD) represents one of the most common conditions at old age, an interplay between genetic factors for AD and longevity is expected.

We explored this interplay by studying the prevalence of 38 AD-associated single-nucleotide-polymorphisms (SNPs) identified in AD-GWAS, in self-reported cognitively healthy centenarians, and we replicated findings in the largest GWAS on parental-longevity.

We found that 28/38 SNPs identified to associate with increased AD-risk also associated with decreased odds of longevity. For each SNP, we express the imbalance between AD- and longevity-risk as an effect-size distribution. When grouping the SNPs based on these distributions, we found three groups: 17 variants increased AD-risk more than they decreased the risk of longevity (AD-group): these variants were functionally enriched for *β*-amyloid metabolism and immune signaling, and they were enriched in microglia. 11 variants reported a larger effect on longevity as compared to their AD-effect (Longevity-group): these variants were enriched for endocytosis/immune signaling, and at the cell-type level were enriched in microglia and endothelial cells. Next to AD, these variants were previously associated with other aging-related diseases, including cardiovascular and autoimmune diseases, and cancer. Unexpectedly, 10 variants associated with an increased risk of both AD and longevity (Unexpected-group). The effect of the SNPs in AD- and Longevity-groups replicated in the largest GWAS on parental-longevity, while the effects on longevity of the SNPs in the Unexpected-group could not be replicated, suggesting that these effects may not be robust across different studies.

Our study shows that some AD-associated variants negatively affect longevity primarily by their increased risk of AD, while other variants negatively affect longevity through an increased risk of multiple age-related diseases, including AD.

## Introduction

The human lifespan is determined by a beneficial combination of environmental and genetic factors.^1,2^ Long-lived individuals tend to cluster in families, suggesting that the role of the genetic factors is considerable,^3,4^ however, the research of genetic variants that influence human lifespan has yielded contrasting results: only the longevity-association of the *APOE* alleles and few additional variants consistently replicated across studies (*CDKN2B, ABO*).^5,6^ While the replication rate in independent studies is low, a large collection of genetic variants has been associated with longevity through genome-wide association studies (GWAS).^5,6^ The majority of these variants was previously identified to associate with other age-related conditions, including cardiovascular disease, autoimmune and neurological disorders, suggesting that the genetics underlying human longevity depends on a low risk for several age-related diseases.^2,5,6^

Of all age-related diseases, late-onset Alzheimer’s Disease (AD) is the most common type of dementia and one of the most prevalent causes of death at old age.^7^ The largest risk factor for AD is aging: at 100 years of age, the disease’s incidence is about 40% per year.^8^ Genetic factors play a significant role in AD as heritability was estimated to be 60–80%:^9^ the strongest common genetic risk factor for AD is the *APOE*-*ε4* allele, and large collaborative GWAS have identified ∼40 additional common variants associated with a slight modification of the risk of AD.^10–13^ Despite high incidence rates of AD at very old ages, AD is not an inevitable consequence of aging, as demonstrated by individuals who surpass the age of 100 years with high levels of cognitive health.^14^

As AD-associated variants increase the risk of AD, leading to earlier death, a negative effect on longevity for these variants is to be expected. However, apart from *APOE* alleles, genetic variants that influence the risk of AD were not found to affect the human lifespan in previous GWAS. When assuming that AD-associated variants affect AD only and that these variants’ effect is constant during aging, then the effect on longevity for these variants should be proportional to their effect on AD, albeit in a different direction. In other words, if a variant increases the risk of AD 2-fold, then carriers will have twice as much AD-related mortality as non-carriers, and as a consequence (given the assumptions), they will have twice as little chance to age into a cognitively healthy centenarian. For example, variant *rs72824905* (Pro522Arg) in the *PLCG2* gene was recently found to decrease the risk of AD 1.75-fold, 1.63-fold frontotemporal dementia, and 1.85-fold dementia with Lewy bodies, while being associated with a 1.49-fold increased likelihood of longevity.^15^ For a variant that is protective against multiple conditions, it might be expected that the overall effect on longevity should be larger than the inverse of the effect on AD alone.

We have previously shown that cognitively healthy centenarians are depleted with genetic variants that increased the risk of AD compared to a general population; however, the extent of depletion was variant specific, suggesting that a subset of AD-variants may be specifically beneficial to reach extremely old ages in good cognitive health.^16,17^ There is, however, little evidence of an age-dependent effect for AD variants and the extent to which these variants affect other age-related diseases is mostly unknown.^18^ Using the assumption of effect-size proportionality, we set out to investigate the relationship between AD- and longevity-risk for genetic variants associated with AD.

## Methods

### Populations and selection of genetic variants

We included N=358 centenarians from the 100-plus Study cohort, which comprises Dutch-speaking individuals aged 100 years or older who self-report to be cognitively healthy, which is confirmed by a proxy.^14^ As population controls, we used population-matched, cognitively healthy individuals from five studies: (*i*) the Longitudinal Aging Study of Amsterdam (LASA, N=1,779),^19,20^ (*ii*) the memory clinic of the Alzheimer center Amsterdam and SCIENCe project (N=1,206),^21,22^ (*iii*) the Netherlands Brain Bank (N=40),^23^ (*iv*) the twin study of Amsterdam (N=201)^24^ and (*v*) the 100-plus Study (partners of centenarian’s children, N=86).^14^ See *Supplementary Methods: Populations* for a detailed description of these cohorts. Throughout the manuscript, we will refer to the union of the individuals from these five studies as population subjects. The Medical Ethics Committee of the Amsterdam UMC (METC) approved all studies. All participants and/or their legal representatives provided written informed consent for participation in clinical and genetic studies.

Genetic variants in our populations were determined by standard genotyping and imputation methods. After establishing quality control of the genetic data (see *Supplementary Methods: Quality control*), 2,905 population subjects and 343 cognitively healthy centenarians were left for the analyses (*Table 1*). We then selected 41 variants representing the current genetic landscape of AD (*Table S1*).^13^ We restricted our analysis to high-quality variants with a minor allele frequency >1% in our cohorts, which led to the exclusion of 3/41 variants (rare variants in the *TREM2* gene *rs143332484* and *rs75932628* and *ABI3* gene *rs616338*), leaving 38 variants for the analyses.

**Table 1:** Population characteristics.

|  | Population controls | Cognitively healthy centenarians |
| --- | --- | --- |
| **Number of individuals** | 2,905 | 343 |
| **Females (%)** | 1400 (48.2) | 246 (71.7) |
| **Age (SD)<sup>a</sup>** | 68.3 (11.5) | 101.4 (1.8) |
| **ApoE ε4 (%)** | 1,012 (17.38) | 48 (7.15) |
| **ApoE ε2 (%)** | 523 (9.00) | 91 (13.26) |

<sup>a</sup> Age at study inclusion; SD, standard deviation; *ApoE*, Apolipoprotein E allele count for ε4 and ε2, and relative allele frequency in population controls and cognitively healthy centenarians. Reference to the cohorts reported in this table are: <sup>14,20-24</sup>

### AD and longevity variant effect sizes

We first retrieved the effect-size on 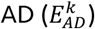 for each AD variant, *k*, from a large genome-wide meta-analysis of AD.^13^ To estimate a confidence interval, we bootstrapped the published effect-sizes (log of odds ratios) and their respective standard errors (*B*=10,000).

To calculate the effect-size on longevity 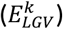 for the same variants, we used a logistic regression model with cognitive healthy centenarians as cases and population subjects as controls while adjusting for population stratification (PC 1-5). The number of principal components to include as covariates was arbitrarily chosen; however, as all individuals were population-matched, we expected these components to correct all major population effects. The resulting *p-values* were corrected for multiple testing (False Discovery Rate, FDR). To calculate the confidence interval, we repeated this procedure for bootstraps (*B*=10,000) of the data. For convenience, variant effect-sizes on AD and longevity were calculated with respect to the allele that increases the risk of AD, such that 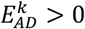.

Given a variant *k*, with a relative effect-size on 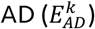 and on longevity 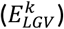, we defined that the variant has an *expected direction* if the variant increases the risk of AD, *i*.*e*. 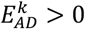, and at the same time decreases the risk of longevity, *i*.*e*. 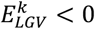. Inversely, we define that the longevity effect has an *unexpected direction* if the allele that increased AD risk also increased the risk of longevity, *i*.*e*. 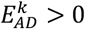 and 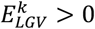. The probability of observing an *expected direction* was considered a Bernoulli variable with *p*=0.5 (*i*.*e*. equal chance of having an *expected/unexpected* direction), thus the number of variants with an *expected direction* follows a binomial distribution.

### Imbalance of variant effect direction

We represented each variant as a data point whose coordinates were defined by the variant’s effect on AD (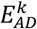, on the y-axis) and its effect on longevity (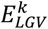, on the x-axis). See *Figure S1* for an example. For each variant, we then calculated the normalized angle, *α*_*k*_, of the vector representing the data point with the x-axis: 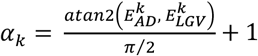, with *α*_*k*_ ∈ [−1; 1]. This normalized angle relates to the imbalance between the risk of AD and the risk of longevity. That is, for *α*_*k*_ < 0 the variant has an expected direction, while for *α*_*k*_ > 0 the variant has an unexpected direction.

As the effect-sizes are sample estimates, we subsequently took their confidence interval into account to create, for each variant, a distribution of the imbalance in the effect direction (*IED*). Hereto, we assumed a Gaussian density for both 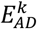 and 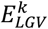, centered around 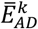 and 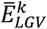 and with a variance equal to the estimated confidence interval for both effect sizes, respectively. We sampled 10,000 times from these distributions and calculated the corresponding imbalance (*α*_*k*_), to get a (non-Gaussian) distribution of the *IED* for that variant, *IED*_*k*_. To group variants with similar patterns of their *IED* distributions, we ordered the *IED* by their median value 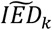, and defined a group of variants in which the effect sizes were in the expected direction 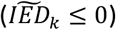, which we subsequently split in those that have (*i*) a larger effect on longevity as compared to the effect on AD (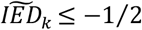, *Longevity-group*), and those that have (ii) a larger effect on AD as compared to the effect on longevity (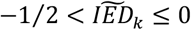, *AD-group*). We defined a third group of variants that have an effect in the unexpected direction (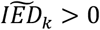, *Unexpected*-*group*).

### Linking variants with functional clusters

To investigate each variant’s functional consequences, we calculated the variant-pathway mapping, which indicates the degree of involvement of each genetic variant in AD-associated pathways (*Figure S2*). See *Supplementary Methods: variant-pathway mapping* for a detailed explanation of our approach. Briefly, the *variant-pathway mapping* depends on (*i*) the number of genes each variant was associated with and (ii) the biological pathways each gene was associated with. We calculated the variant-pathway mapping for all 38 AD-associated variants. Finally, we compared the variant-pathway mapping within each group of variants defined based on the *IEDs* (Longevity-, AD- and Unexpected-groups) using Wilcoxon sum rank tests and correcting *p-values* using FDR: this was indicative of whether a group of variants was enriched for a specific functional cluster (*Figure S2*).

### Cell-type annotation at the level of each cluster

To further explore the biological basis of the different groups of variants (Longevity-, AD- and Unexpected-groups), we calculated the degree of enrichment of each group for specific brain cell-types (see *Supplementary Methods: cell-type annotation* for a detailed description). This annotation depends on the number of genes each variant was associated with, and the expression of these genes in the different brain cell-types, *i*.*e*. astrocytes, oligodendrocytes, microglia, endothelial cells, and neurons. We finally compared the cell-specific annotations within each group of variants (Longevity-, AD- and Unexpected-groups) using Wilcoxon sum rank tests and correcting *p-values* using FDR, which indicated whether a group of variants was enriched for specific brain cell-types (*Figure S2*).

### Replication of findings in large GWAS cohorts

To find additional evidence for our findings, we inspected the association statistics of the 38 AD-associated variants in the largest GWAS on parental longevity.^6^ Briefly, in this study offspring’s genotypes were used to model parental age at death. In this dataset, we looked at the significance of association with longevity for the 38 variants (*p-values* were corrected with FDR) and their direction of effect. Finally, we tested the consistency in the expected/unexpected directions between our study and the GWAS on parental longevity using binomial tests.

### Implementation

Quality control of genotype data, population stratification analysis and relatedness analysis were performed with PLINK (v2.0 and v1.9). All subsequent analyses were performed with R (v3.6.3), Bash, and Python (v3.6) scripts. All scripts are freely available at https://github.com/TesiNicco/Disentangle_AD_Age. Variant-gene annotation and gene-set enrichment analyses are implemented in a stand-alone package available at https://github.com/TesiNicco/AnnotateMe. Annotation and gene-set enrichment analysis of SNP-sets can also be run on our web-server at https://snpxplorer.eu.ngrok.io.

## Results

### AD-associated variants also associate with longevity

We explored the association with longevity of 38 genetic variants previously associated with AD in GWAS (*Table S1*). We tested these variants in 343 centenarians who self-reported to be cognitively healthy (mean age at inclusion 101.4±1.3, 74.7% females), as opposed to 2,905 population subjects (mean age at inclusion 68.3±11.5, 50.7% females). We found a significant association with longevity for two variants after multiple testing correction (FDR<5%, variants in the *APOE* gene; *rs429358* and *rs7412, Table S2*). We compared the direction of effect on longevity with that on AD as found in literature: of the 38 variants, 28 showed an association in the expected direction, *i*.*e*. alleles that increased AD risk were associated with lower odds of longevity, and this was significantly more than expected by chance (*p*=0.005 including *APOE* variants, *p*=0.01 excluding *APOE* variants, see *Methods*).

### Distributions of the imbalance in the effect direction (*IED*)

To study the relationship between the effect on AD and longevity for all 38 AD-associated variants in more detail, we created distributions of the imbalance in the variant effect direction (*IED*): *Figure 1*. The *IED* of a variant indicates whether the effects on AD and longevity are in the expected direction (values<0) or in the unexpected direction (values>0). A variant with an *IED* value approaching 0 has a pure AD effect (AD-end), while a variant with values *IED* value close to -1 has a pure longevity effect (Longevity-end); see *Methods* for a detailed explanation. Based on the median value of each *IED* distributions, 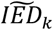, we grouped the variants into (*i*) a Longevity-group (variants with a 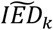 skewed towards the longevity*-*end of the spectrum), (*ii*) an AD-group (variants with a 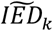 skewed towards the *AD*-end of the spectrum), and (*iii*) an Unexpected-group (variants with a 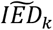 in the unexpected direction). The *AD-*group included 17 variants (*APOE* (1), *APOE* (2), *SCIMP, PLCG2* (1), *MS4A6A, BIN1, PILRA, APP, PLCG2* (2), *CR1, SLC24A4, TREML2, ACE, APH1B, FERMT2, PICALM, CD33*) and the longevity*-*group included 11 variants (*SHARPIN* (1), *SHARPIN* (2), *HS3ST1, EPAH1, IQCK, PRKD3, CD2AP, PLCG2* (3), *SPI1, HLA, EDHDC3*), such that the effect of 28/38 (74%) of all variants was in the expected direction. The effect of 10 variants was in the unexpected direction, the Unexpected-group: (*PTK2B, CLU, KANSL1, INPP5D, ABCA7, CHRNE, SORL1, IL34, ADAM10, CASS4*) (*Figure 1*).

**Figure 1:**
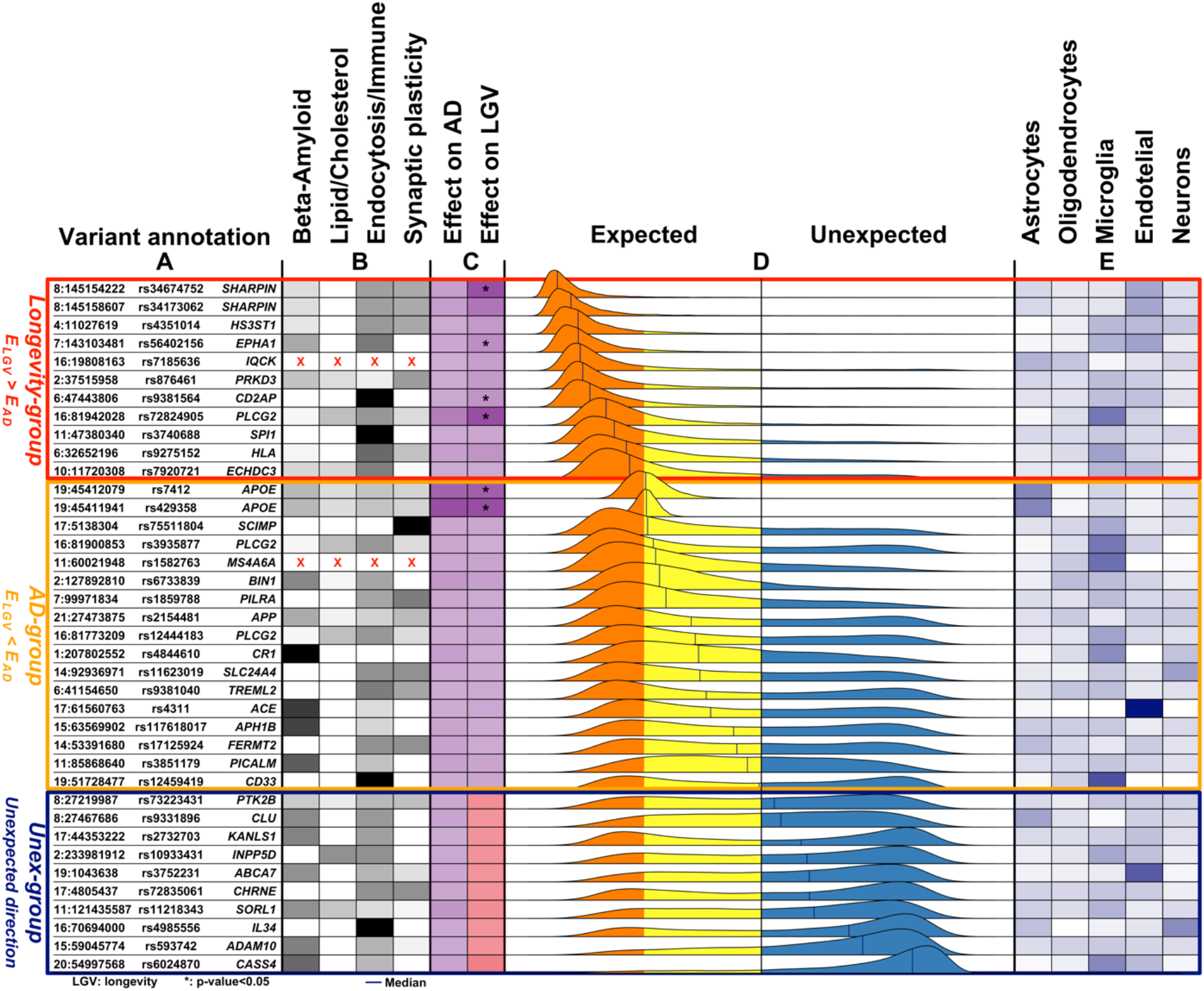
Overview of the 38 genetic variants associated with Alzheimer’s disease. **A**. The genomic position of the variants (chromosome: position), variant identifier, and closest gene. Genomic positions are with respect to GRCh37 (hg19). **B**. The variant-pathway mapping score of association with the four functional clusters (darker colors representing stronger associations). Variants annotated with red crosses could not be annotated to any one of the functional clusters as no biological processes are associated with the related genes. **C**. The effect size on AD (from literature) and the observed effect size on longevity (LGV) for each variant (darker color indicating stronger effect). The same color indicates *expected direction* (*i*.*e*. increased risk of AD and decreased chance of longevity), while different colors, visible in the *Unex-group* of variants, indicates *unexpected direction*. For the longevity effects, we also annotate variants for which we observed a significant association (unadjusted *p-value*<0.05). **D**. The distribution of the imbalance direction of variant effect (*IED*) in AD-risk as compared to cognitive health aging (see *Methods* for details). The *longevity-, AD-* and *Unex-groups* were derived based on the median value of the *IED*. The median value is reported for each *IED* as a blue vertical line. **E**. Average gene expression of the genes associated with the variant in five different brain cell-types (the darker, the higher the expression).

### AD-associated variants in large GWAS of longevity

To find additional evidence for longevity associations, we inspected the AD-associated variants’ effect in the largest GWAS on parental longevity.^6^ Of the 38 AD-associated variants, association statistics were available for 34 of the variants (missing from longevity-group: *PLCG2* (3), *SPI1;* missing from Unexpected-group: *KANSL1, INPP5D*). Overall, 21/26 (81%) of the variants in the expected direction in our study (of which 6/9 variants in Longevity- and 15/17 variants in the AD-group), were also in the expected direction in the independent parental longevity dataset. Variants in the expected direction in the first analysis are significantly more likely to be in the expected direction in the replication analysis than in the unexpected direction (*p*=0.01, based on a binomial test, *Figure 3*). Six AD-associated variants reached significance in the parental-longevity GWAS after correcting for multiple comparisons (FDR<5%): variants in the *APOE* gene (*rs429358* and *rs7412)* and variants in/near *PRKD3 (rs8764613), CD2AP (rs9381564), APH1B* (*rs117618017)* and *BIN1 (rs6733839*). Of these, variants in/near *PRKD3* and *CD2AP* belonged to the Longevity-group in our analysis.

**Figure 3:**
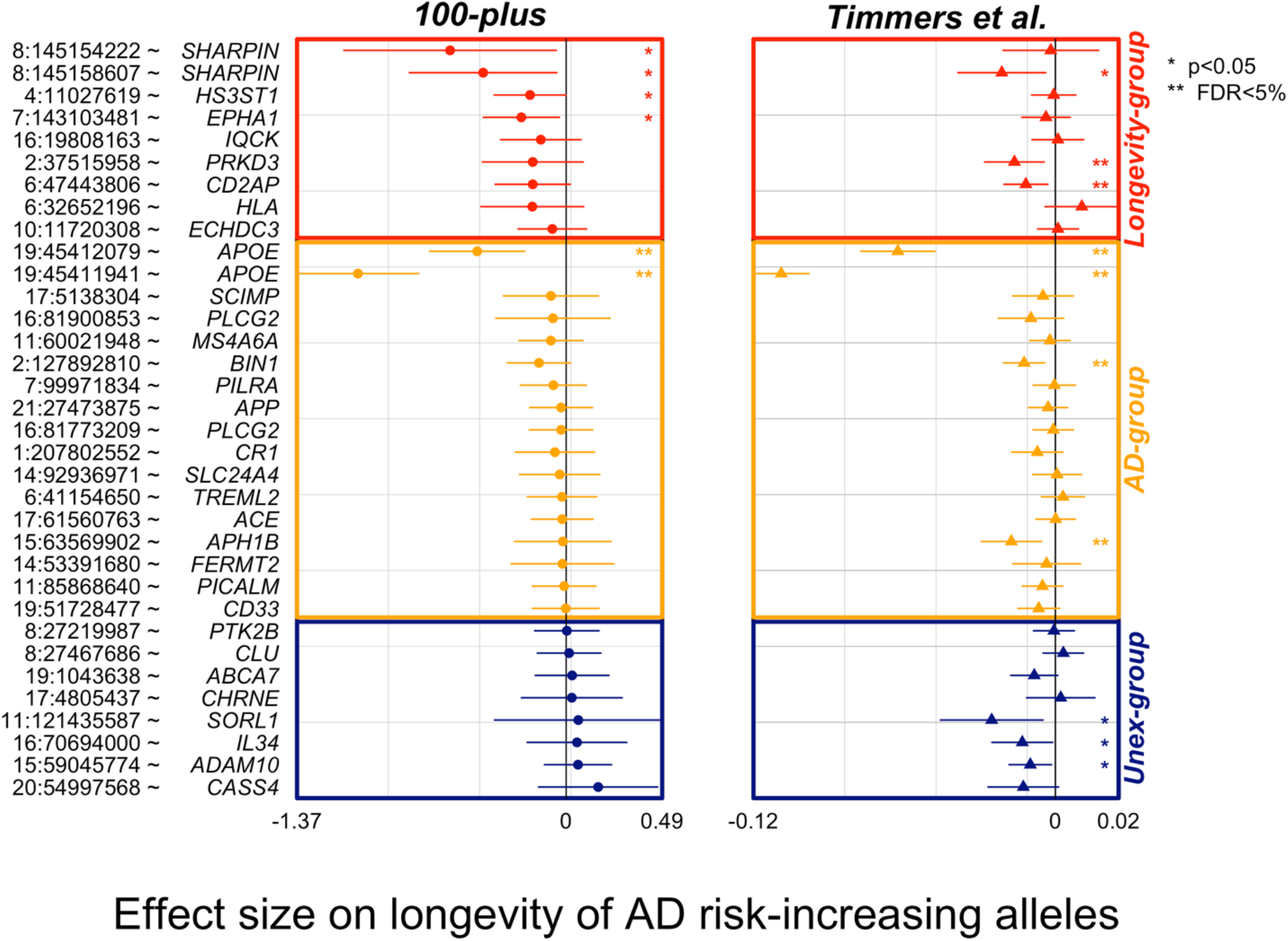
Forest plot of association statistics of AD-variants in our study and the largest GWAS of parental longevity. The plot shows the association of AD-variants in our study and the largest by-proxy GWAS on parental longevity.^6^ The association statistics of 34/38 variants were available from publicly available summary statistics of Timmers et al. study. Plotted effect-sizes are with respect to the AD-risk increasing allele. Thus, an expected direction of effect is shown for variants with a negative estimate. Nominally significant associations with AD (*p<*0.05) are annotated with an asterisk (*), and significant associations after FDR correction are annotated with two asterisks (**).

Conversely, only 2/8 (25%) variants that we observed in the unexpected direction in our study were also in the unexpected direction in the parental-longevity GWAS, such that these variants were *not* more likely to be in the unexpected direction (*p=*0.29, based on a binomial test, *Figure 3*).

### Functional characterization of variants

The 38 AD-associated variants included coding variants (N=10), intronic variants (N=20), and intergenic variants (N=8) (*Table S3*). 12/28 of the intronic/intergenic variants had eQTL associations. In total, the 38 variants mapped to 68 unique genes, with most variants mapping to one gene (N=21) and fewer mapping to 2 genes (N=10), 3 genes (N=2), 4 genes (N=1), 5 genes (N=2), 6 and 7 genes (N=1, respectively) (*Figure S3* and *Table S3*).

We performed gene-set enrichment analysis using a sampling-based approach to explore the biological processes enriched in the 68 genes associated with AD-variants (see *Methods* and *Figure S2*). We found 115 significantly enriched biological processes after correction for multiple tests (FDR<5%, *Table S4*). After clustering these terms based on their semantic similarity, we found four main clusters of biological processes: (*i*) *β*-amyloid metabolism, (*ii*) lipid/cholesterol metabolism, (*iii*) endocytosis/immune signaling and (*iv*) synaptic plasticity (*Figure 1, Figure S4* and *Table S5*).

Next, we calculated the *variant-pathway mapping score* (see *Methods* and *Figure S2*), which indicates how well a variant is associated with each of the 4 functional clusters. In total, we calculated the *variant-pathway mapping* for 30 variants; we imputed the annotation of 6 variants (*Table S5* and *Table S5*), while 2 variants could not be annotated (variants *rs7185636* and *rs1582763* in/near *IQCK* and *MS4A6A* genes), because the associated genes were not annotated with any biological process function (*Table S5*). Finally, we tested whether the Longevity-, AD- and Unexpected-groups were enriched for specific functional clusters by comparing the distribution of variant-pathway mapping within each group (see *Methods, Figure 2*, and *Figure S2*). The Longevity-group was significantly (FDR<10%) enriched for the endocytosis/immune signaling functional cluster; the AD-group for the endocytosis/immune signaling, *β* -amyloid metabolism and to a smaller extent for the synaptic plasticity functional clusters; the Unexpected-group was mainly enriched for the endocytosis and *β*-amyloid metabolism functional clusters.

**Figure 2:**
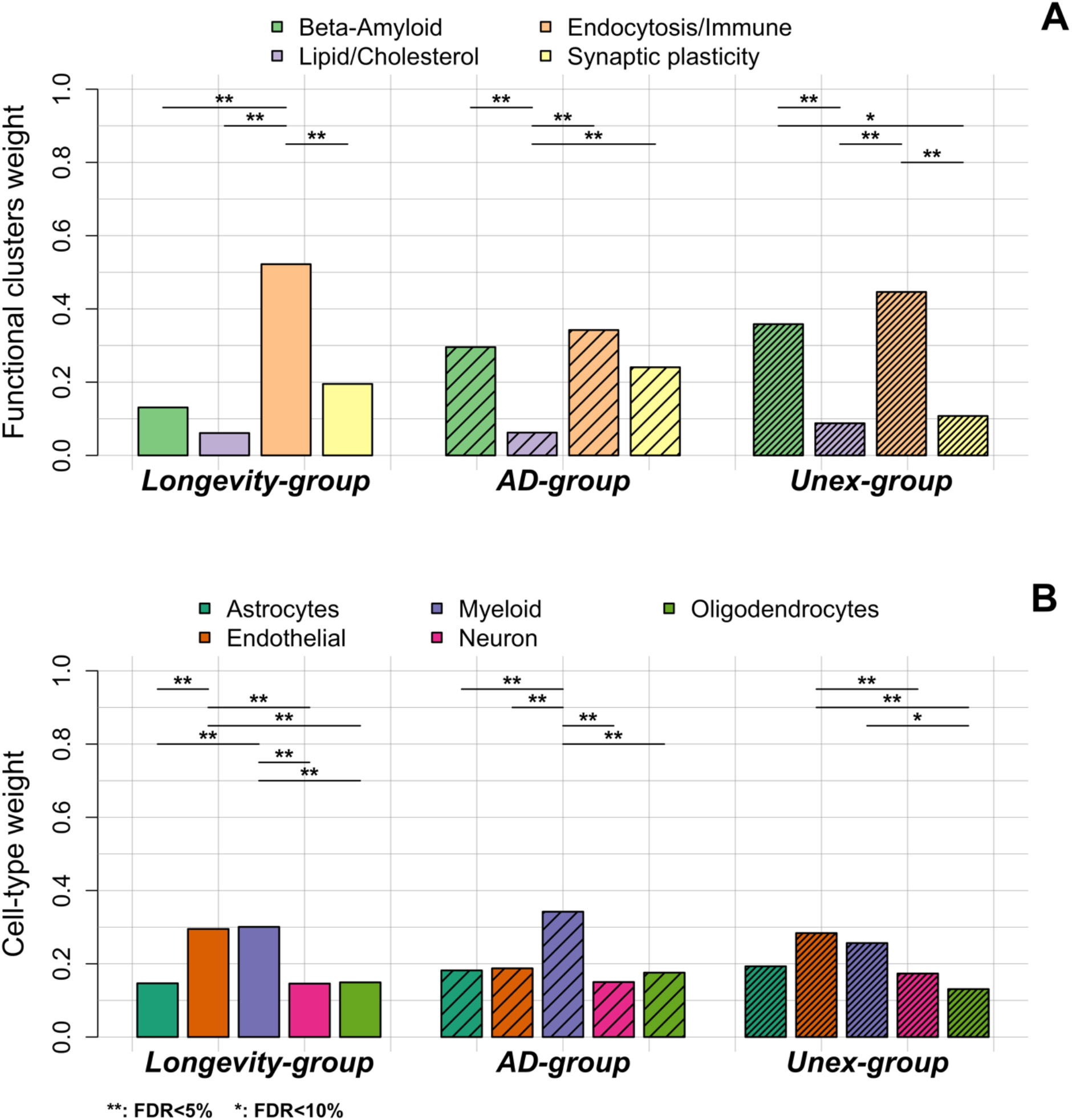
Comparison of functional annotation and cell-type annotation within the *Longevity-, AD-* and *Unex-groups*. **A**. The weights of the 4 functional clusters within the *longevity-, AD-* and *Unex-groups*. **B**. The weights of the different cell-types in the brain, per group. Differences in functional weights and cell-type weights within each group were calculated using Wilcoxon sum rank tests. The resulting p-values were FDR-corrected.

### Expression of AD-associated genes in brain cell-types

We explored whether specific brain cell types, *i*.*e*. astrocytes, oligodendrocytes, microglia, endothelial cells and neurons, were enriched within each group of variants (see *Methods, Table S5*, and *Table S6*). *Figure 1* shows the collapsed cell-type expression for all 38 AD-associated variants. We then tested the enrichment for cell-type expression within the Longevity-, AD- and Unexpected-groups. The Longevity*-*group was significantly enriched for myeloid and endothelial cells, the *AD-*group for myeloid cells, while the Unexpected*-*group was significantly enriched for endothelial cells (FDR<10%).

## Discussion

### Summary of the findings

We studied the effect on longevity of 38 genetic variants previously associated with AD through GWAS.^13^ We found that a majority of 74% of the alleles that increase the risk of AD is associated with lower odds of becoming a centenarian (expected direction). Overall, most variants (N=17) had a larger effect on AD than on longevity: these variants were associated with *β* -amyloid metabolism and endocytosis/immune signaling, and were primarily expressed in microglia. A subset of variants (N=11) had a larger effect on longevity than their effect on AD. These variants were associated mostly with endocytosis and immune signaling, and they were expressed in microglia and endothelial cells. These variant-effects were confirmed for 81% of the alleles in an independent dataset, the largest GWAS on parental longevity. In contrast, 26% of the variants increased both the risk of developing AD *and* the risk of becoming a centenarian (n=10), (unexpected direction). These unexpected effects could only be replicated for 2 of the variants in the independent dataset, suggesting that the expected effects were more robust across studies than the unexpected effects. Together, our findings suggest that variants associated with AD-risk may also be linked with the risk of other age-related diseases, and that survival/longevity is affected by these variants.

### AD-associated variants and their effect on healthy aging

A single study previously explored the extent to which 10 AD-associated variants affect longevity: apart from *APOE* locus, none of the other 10 tested AD-associated variants significantly associated with longevity.^25^ In addition to *APOE*, four variants showed a negative effect on longevity while increasing AD-risk (in/near *ABCA7, EPHA1, CD2AP*, and *CLU*). In agreement with these findings, we also found that only the *APOE* variants significantly associated with longevity, and variants in/near *EPHA1* and *CD2AP* belong to the Longevity-group. However, in our study, we found that most alleles associated with an increased risk of AD associated with a decreased chance of longevity. The inability to observe such an inverse relationship between variant effects on AD and longevity in the previous study may be explained by the relatively small sample sizes, combined with a low number of (well-established) AD variants analyzed (N=10). In our study, groups sizes were also relatively small, but the centenarians had a relatively high level of cognitive health, which might have contributed to an increased effect size of AD-associated genetic variants in our comparison.^16,17^

### Different trajectories of effect of AD-associated variants on healthy aging

#### Variants with a larger effect on AD than longevity

For most variants with effects in the expected direction, the risk-increasing effect on AD was more extensive than the negative effect on survival/longevity. These variants, which include both *APOE* alleles, might negatively affect lifespan because carriers are removed from the population with increasing age due to AD-associated mortality. For the *APOE* variants specifically, the distribution of the imbalance in the effect directions suggests a nearly similar proportion of the increased risk of AD and decreased risk of longevity for both *APOE* variants 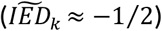. This explains why multiple previous studies have associated *APOE* variants with longevity. In our cohort of centenarians, the frequency of the deleterious *ε4* allele is half of that of the population controls (8% *vs*. 16%, respectively). In comparison, the frequency of the protective *ε2* allele is nearly two-fold increased (16% *vs*. 9%).^16^ Note, however, that inclusion criteria of the centenarian cohort required them to self-report to be cognitively healthy, which might have increased the observed longevity effect. Apart from the *APOE* variants, the AD-group included 15 variants, all of which were among the first to be associated with AD through GWAS (*CR1, CD33, BIN1, MS4A6A, PICALM*, and *SLC24A4*).^26,27^ These common variants have the strongest associations with AD: largest odds ratios (OR) leading to lowest *p*-values. Functional annotation showed significant enrichment of *β*-amyloid metabolism, which aligns with the importance of functional APP metabolism in maintaining brain health. We also observed functional enrichment of endocytosis and immune signaling, and a specific cell-type enrichment for microglia. This is in line with the currently growing hypothesis of the involvement of immune dysfunction in the etiology underlying AD.^28,29^

#### Variants with a larger effect on longevity than AD

The second-largest group of variants constituted a subset of 11 variants with a larger effect on longevity than the effect on AD, which suggests that these variants may be involved in other age-related diseases or general age-related processes. The AD-association of most of these variants is relatively recent, likely due to small effect sizes (ORs) or variants rareness (low minor allele frequency, MAF); both features necessitate a very large number of samples to identify these variants as significantly associating with the disease. The variants within this group were specifically enriched for immune response and endocytosis, which are known hallmarks of longevity.^1,30,31^ In addition to the rare non-synonymous variant in the *PLCG2* gene (*rs72824905*, MAF: 0.6%), which was recently observed to be protective against AD, frontotemporal dementia (FTD) and dementia with Lewy bodies, other variants within this group were previously linked with disease risk factors. One of the two non-synonymous variants in the *SHARPIN* gene, variant *rs34173062* (MAF: 5.7%), has been associated with respiratory system diseases in GWAS.^32–34^ Variant *rs7185636* (MAF: 17.1%), intronic of the *IQCK* gene, is in complete linkage with a variant (*rs7191155*, R^2^=0.95), which was previously associated with body-mass index (BMI).^35^ The variant *rs876461* (MAF: 13.0%) near the *PRKD3* gene is in linkage with variant *rs13420463* (R^2^=0.42), which has been associated with systolic blood pressure.^36^ Further, the variant near *CD2AP* gene associates with the development and maintenance of the blood-brain barrier, a specialized vascular structure of the central nervous system which, when disrupted, has been linked with epilepsy, stroke and AD.^37^ Variant *rs9275152* (MAF: 10.4%) maps to the complex Human-Leukocyte-Antigen (HLA) region, which codes for cell-surface proteins responsible for the regulation of the adaptive immune system. In numerous GWAS, variants in the HLA region were associated with autoimmune diseases, cancer, and longevity.^6,38^ The AD-associated variant in this region (*rs9275152*) is also a risk variant for Parkinson’s disease.^39^ Finally, the genomic region surrounding the *SPI1* gene (in which variant *rs3740688* maps) has been previously associated with cognitive traits (intelligence, depression)^40(p300)^ and, with lower evidence, with kidney disease and cancer.^41,42^ The remaining variants *rs56402156, rs7920721*, and *rs4351014* (in/near *EPHA1, ECHDC3*, and *HS3ST1*) have not been directly associated with other traits, although their associated genes were implicated in systemic lupus erythematosus (*HS3ST1*) and cancer (*EPHA1, ECHDC3*).^43,44(p1),45^ Together, these findings suggest that the counterpart of each risk-increasing allele, the AD-protective alleles, might give a survival advantage that is not only specific to AD. Their functional and cell-type annotations suggest that they contribute to the maintenance of regulatory stimuli in the immune and endosomal systems, which may be essential to maintain brain and overall physical health, necessary to reach extremely old ages in good cognitive health.^17^

### Variants associated with increased risk of AD and increased longevity risk: unexpected group

Unexpectedly, ten variants increased the risk of AD while at the same time increasing the chance to reach ages over 100 in good cognitive health, which is an *unexpected* balance. We note that the *IED* distributions of these variants were broad, and in some cases even showed a bimodal behavior (in/near *KANSL1, IL34, CHRNE*): this is attributable to the small effect-sizes (and large standard errors) on longevity for these variants, which caused data points to easily flip between the expected and unexpected direction during the sampling procedure. Replication of the direction of the variant effect in an independent dataset of parental longevity indicated that the unexpected direction was replicated in only the *CLU* and *CHRNE* variants, suggesting that future studies will have to further explore (the robustness of) these unexpected effects.

One explanation for such counter-intuitive effects may be a variant interaction with other variants, which was shown for the variant in the *KANSL1* and *CLU* gene with respect to the *APOE* genotype.^46^ Therefore, carrying the risk allele of such variants may specifically affect the risk of AD in *APOE ε4* allele carriers, which are not prevalent among cognitively healthy centenarians.

An alternative explanation may be that these variants have age-dependent effects: for example, high blood pressure at midlife increases the risk of AD, but after the age of 85 a high blood pressure protects against AD.^47^ Similarly, a high body-mass-index (BMI) increases the risk of AD at midlife, while being protective at older ages.^48^ In line with this hypothesis, the AD variant in/near *IL34* gene codes for a cytokine that is crucial for the differentiation and the maintenance of microglia.^49^ Although further studies are needed, an excessive differentiation in middle-age individuals may increase brain-related inflammation and AD-risk, while it might compensate for the slower differentiation and immune activity at very old ages. Indeed, next to *IL34*, several genes that may be affected by these Unexpected-variants, such as *PTK2B* and *INPP5D*, play a role in aging-associated processes, such as cellular senescence or immunity.^50,51^

### Strengths and weaknesses

We acknowledge that our findings are based on relatively small sample sizes, especially for the cognitively healthy centenarian group. This phenotype is rare, and individuals need to be individually approached for study inclusion,^14^ which is prohibitive for large sample collection. As population subjects in our comparison, we used individuals from five different cohorts: all from the same (Dutch) population, all tested cognitively intact, and did not convert to dementia at the time of analyses. Analyses of variants with low sample sizes in small samples lead to effect sizes large confidence intervals: we took this uncertainty into account by bootstrapping effect sizes, causing the distributions of the imbalance in the variant effect direction of several variants to be widely spread. Although our work represents a first step towards understanding the effect of AD-associated variants on longevity, similar analyses in larger oldest-old or centenarian samples are necessary to support our findings further. Secondly, we had to deal with the problem in GWAS studies, that the driving effect underlying the AD-association of each variant is unclear. Several genes usually map to a specific GWAS locus, represented by one variant. Therefore, to accommodate this uncertainty, we allowed multiple genes mapping to the GWAS locus to be associated with each variant. The subsequent functional annotation of these genes is mainly dependent on the current (limited) knowledge about variant effect and genes function. It is thus likely that our variant-function annotation will change as we gain more understanding about these variant-gene-effects, as well as annotations of gene-functions. When we inspected the parental-longevity GWAS, most of the variants that were in the *expected direction* in our study were also in the same direction in the GWAS; however, this was not true for all variants. The variant that deviated the most between our study and the parental-longevity GWAS was *rs9275152* in the *HLA* region: while we clustered this variant in the Longevity-group, in the parental-longevity GWAS the direction of effect was opposite (*i*.*e. unexpected*), suggesting that the variant increased the risk of AD and at the same time the chance of a long lifespan.^6^ The genomic region to which *HLA* maps is biologically known to be affected by many recombination events and may be population- and environment-dependent, which may explain this divergence.^52^ In addition to *HLA*-variant, variant *rs34674752* in the *SHARPIN* gene reported the second-largest effect-size in our study (after *APOE-ε4*), while the effect-size of this variant in the GWAS was very small, yet in the *expected direction*. To this end, we note that the individuals used in the parental-longevity GWAS were themselves not extremely old individuals, such that possible pleiotropic effects at very old ages, as described earlier, may not be observable in this GWAS. However, while we observed overall consistency in effect-size direction for variants in the expected direction, 6/8 of the variants in the unexpected direction were in the expected direction in the GWAS, with variants near *SORL1, IL34*, and *ADAM10* having the most noticeable differences. We speculate that the relatively young ages of the GWAS samples, together with the small sample size of our centenarian cohort may be the cause of such discrepancy.

## Conclusions

Each AD-associated variant has a different effect on longevity. Variants that have a larger effect on longevity than on AD were previously associated as disease risk-factors, and associated genes are selectively enriched for endocytosis and immune signaling functions.

## Data Availability

Data availability: data that support the findings of this study are available on request, if reasonable, from the corresponding author. The data are not publicly available due to privacy or ethical restrictions.

## Acknowledgments

The following studies and consortia have contributed to this manuscript. Amsterdam Dementia Cohort (ADC): Research at the Alzheimer center Amsterdam is part of the neurodegeneration research program of Amsterdam Neuroscience. 100-plus Study: we are grateful for the collaborative efforts of all participating centenarians and their family members and/or relatives. Wiesje van der Flier holds the Pasman chair. Longitudinal Aging Study of Amsterdam (LASA): the authors are grateful to all LASA participants, the fieldwork team and all researchers for their ongoing commitment to the study.

## Funding

The Alzheimer center Amsterdam is supported by Stichting Alzheimer Nederland and Stichting VUmc fonds. The clinical database structure was developed with funding from Stichting Dioraphte. The SCIENCe project is supported by a research grant from Gieskes-Strijbis fonds and stichting Dioraphte. Genotyping of the Dutch case-control samples was performed in the context of EADB (European Alzheimer DNA biobank), funded by the JPco-fuND FP-829-029 (ZonMW projectnumber 733051061). The 100-plus Study was supported by Stichting Alzheimer Nederland (WE09.2014-03), Stichting Diorapthe, horstingstuit foundation, Memorabel (ZonMW projectnumber 733050814), and Stichting VUmc fonds. Genotyping of the 100-plus study was performed in the context of EADB (European Alzheimer DNA biobank) funded by the JPco-fuND FP-829-029 (ZonMW projectnumber 733051061). Longitudinal Aging Study Amsterdam (LASA) is largely supported by a grant from the Netherlands Ministry of Health, Welfare and Sports, Directorate of Long-Term Care.

## Author contributions

Conceptualization, N.T., M.Hul., S.L., M.R. and H.H.; Data curation, S.L., M.Hul., I.J., N.S., N.Sc., P.S., W.F., M.Hui. and H.H.; Formal analysis, N.T.; Funding acquisition, P.S., W.F., M.Hui., M.R. and H.H.; Methodology, N.T., M.Hul., S.L., M.R. and H.H.; Project administration, P.S., W.F., M.Hui., M.R. and H.H.; Software, N.T., S.L., M.Hul., M.R.; Supervision, N.T., S.L., M.Hul., M.R. and H.H.; Visualization, N.T.; Writing – original draft, N.T., S.L., M.Hul., M.R. and H.H.; Writing – review & editing, N.T., S.L., M.Hul., I.J., N.S., N.Sc., P.S., W.F., M.Hui., M.R. and H.H.

## Conflicts of Interest

All the authors in the study declared no conflict of interest. The funders had no role in the study’s design at any stage.

## Data availability

data that support the findings of this study are available on request, if reasonable, from the corresponding author. The data are not publicly available due to privacy or ethical restrictions.

---

**Figure S1:**
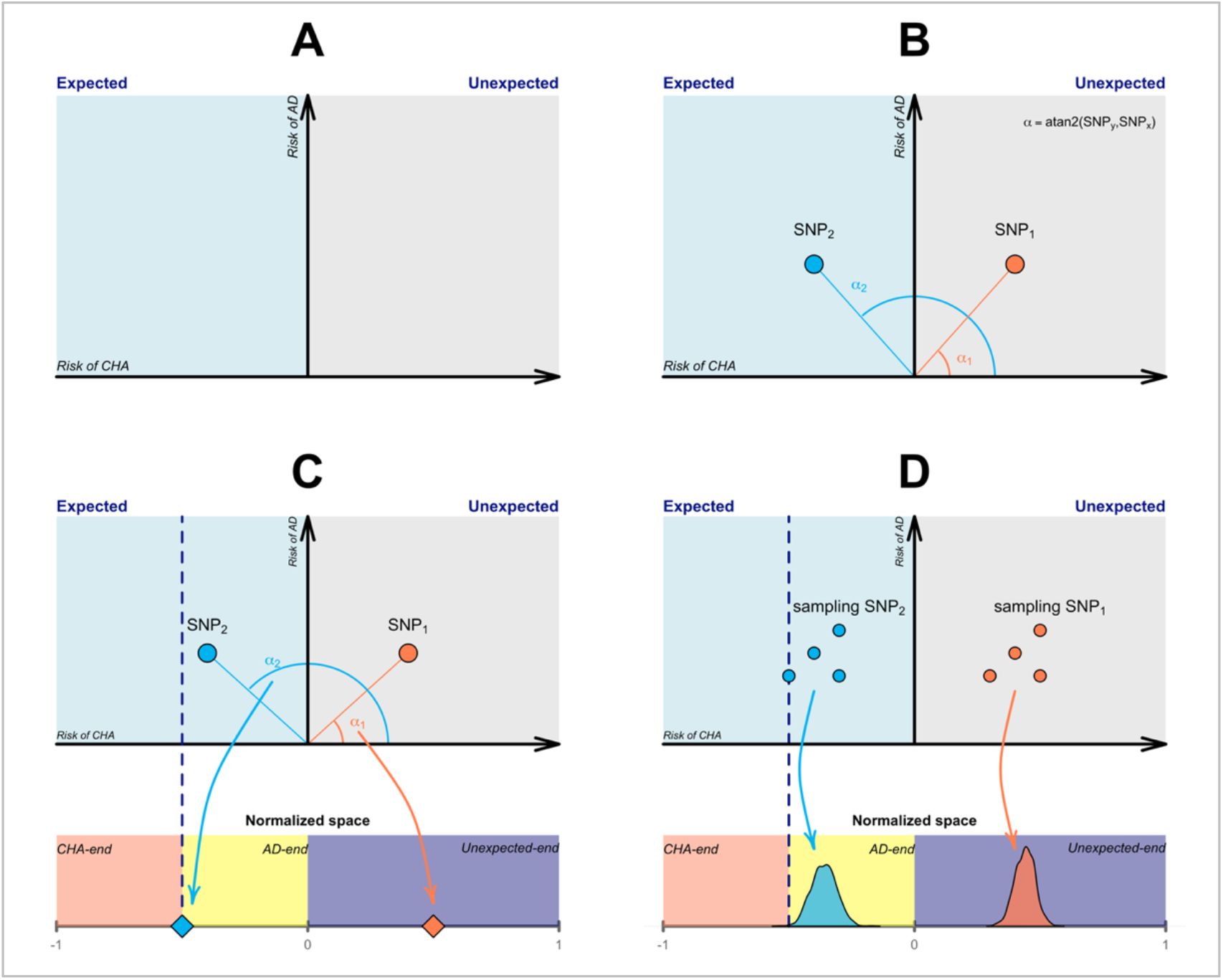
Explanation of the distribution of imbalance variant effect direction (IED). The figure shows the sequential steps for constructing the distribution of the expected direction of variant effect for AD-risk compared to longevity for two toy variants (*SNP*_1_ and *SNP*_2_). **A**. Axes definition, with the y-axis being the effect-size for AD-risk (log of odds ratio) of a variant, derived from literature and set positive by definition. The x-axis identifies the effect-size of a variant on longevity. This can be either positive or negative depending on the variant’s association in cognitively healthy centenarians as opposed to population subjects. The blue area represents that the two effects are in the expected direction with respect to each other, *i*.*e*. a variant increases the risk of AD and at the same time decreases the chance of longevity. Oppositely, the grey area refers to the unexpected direction of effect. **B**. Two toy variants (*SNP*_1_ and *SNP*_2_) are shown as data points. *α*_1−2_ represents the angle of the data point vector with the x-axis. **C**. Normalization of the *α*_1−2_ value into an arbitrary space. Here, we used [−1; 1]. **D**. Repeating this procedure for each bootstrap iteration of each variant, we obtained the distribution of imbalance effect direction for each variant (*IED*). Values smaller than 0 indicate the *expected direction* of effect, whereas values larger than 0 refer to the *unexpected direction* of effects. Additionally, values close to 0 indicate a larger AD effect than longevity effect, and values close to -1 suggest that the variant’s longevity effect is larger than the AD effect.

**Figure S2:**
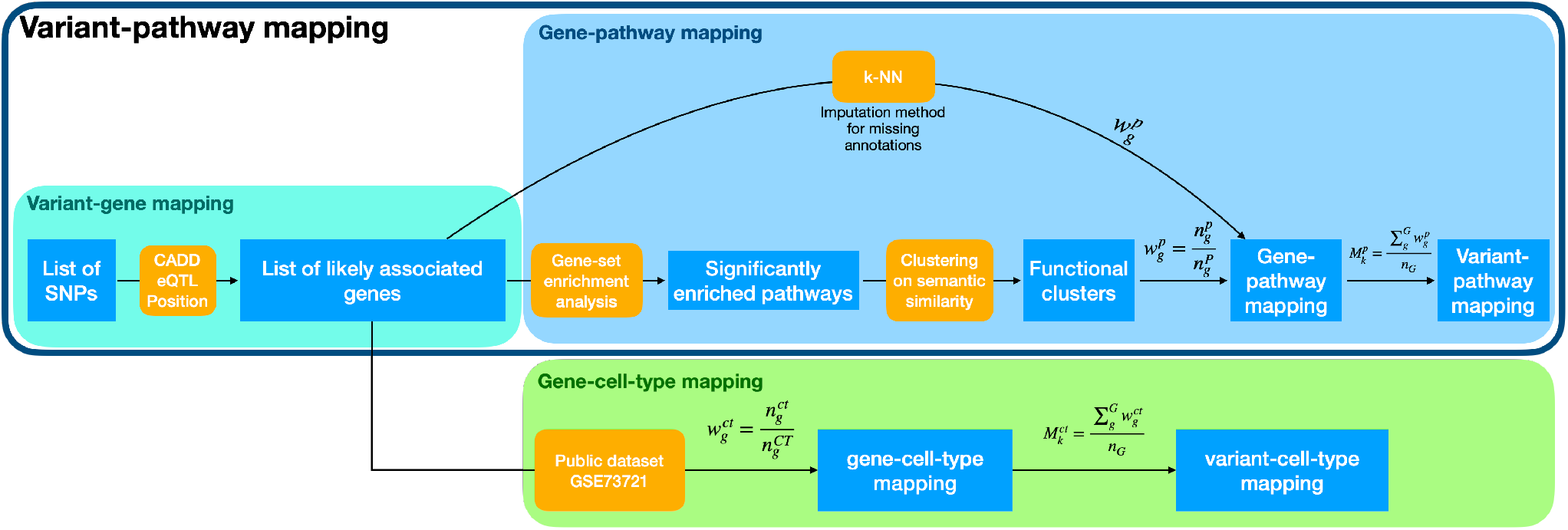
Schematic representation of the variant-pathway and variant-cell-type mapping. The figure shows a schematic representation of the annotation framework used to functionally annotate AD-associated variants and perform cell-type enrichment. Outputs are represented as blue squares, while methods are represented in orange. In the variant-gene mapping, showed in the grey box, we start from a list of variants and, through the integration of predicted variant consequences (CADD), eQTL and position, we obtain a list of genes. Note that here multiple genes may be associated with each variant. The yellow box shows the gene-pathway mapping: briefly, we perform gene-set enrichment analysis followed by clustering of the significantly enriched pathways to obtain functional clusters. We then calculate the gene-pathway mapping by looking at the (enriched) pathways associated with each gene and their associated functional clusters to get a weight for each gene-functional cluster association. Finally, we average the gene-pathway mapping of each gene associated with the same variant. Imputation methods (k-NN) are implemented for genes with missing annotation to obtain the gene-pathway mapping. Together, the grey box and the yellow box form the variant-pathway mapping. At the bottom, the green box shows the gene-cell-type enrichment using the public dataset GSE73721 of gene expression in different brain cell-types. Similar to the gene-pathway mapping, we calculate a weight of association of each gene to each cell-type, and we average these weights in case multiple genes mapped to the same variant (variant-cell-type mapping).

**Figure S3:**
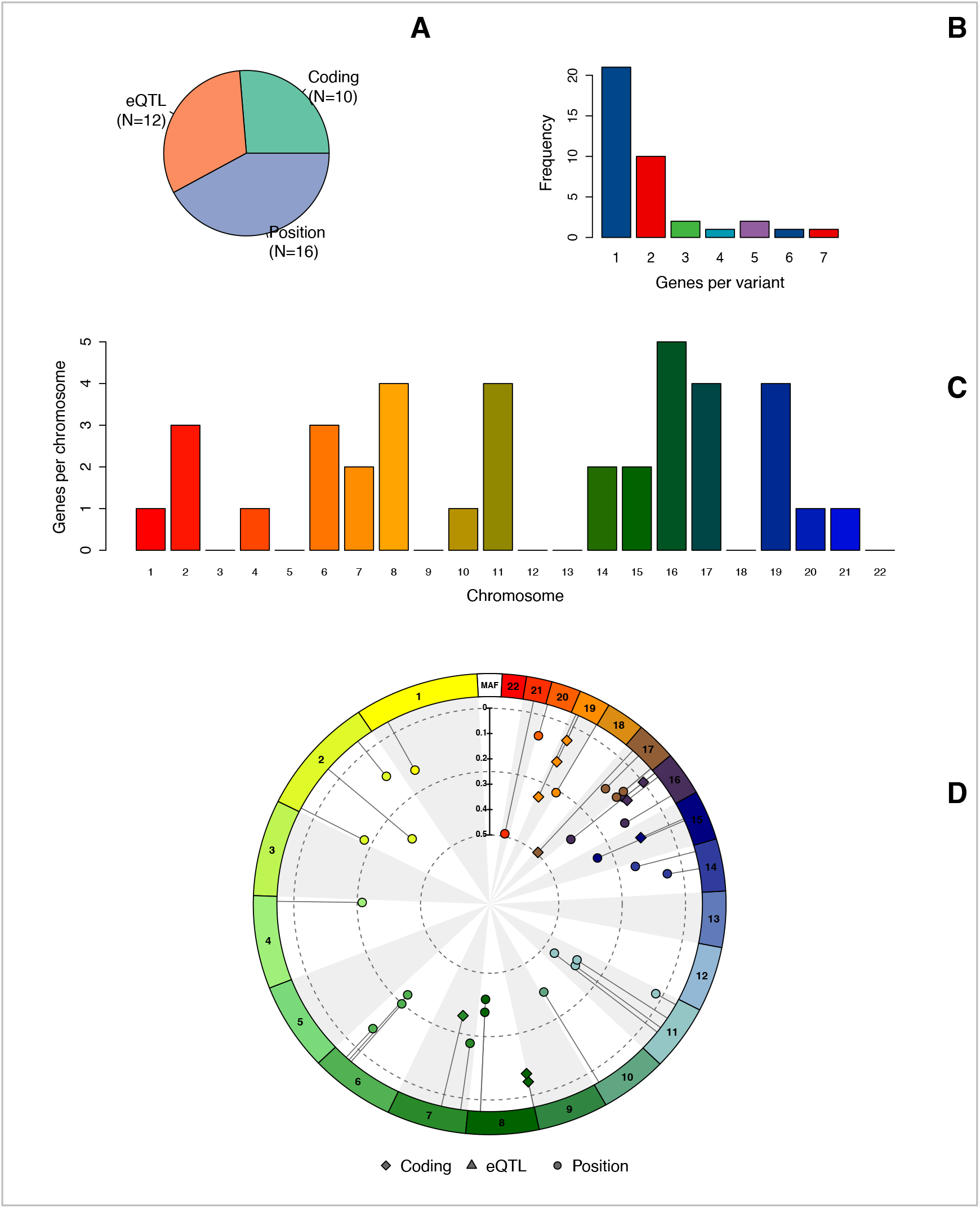
Variant-gene mapping for the 38 AD-associated variants. **A**. The sources used to annotate each variant to the likely affected genes. Coding: variants located in the coding region of a gene (*e*.*g*. synonymous or non-synonymous variants). eQTL: variants associated with RNA expression changes in blood from the GTEx consortium. *Position*: variants intronic or intergenic without evidence of eQTL associations that were annotated based on neighboring genes. **B**. Barplot of the number of genes associated with each variant. **C**. Distribution of genes across the chromosomes. **D**. Distribution of the previously identified variants along the genome together with each variant’s minor allele frequency and annotation.

**Figure S4:**
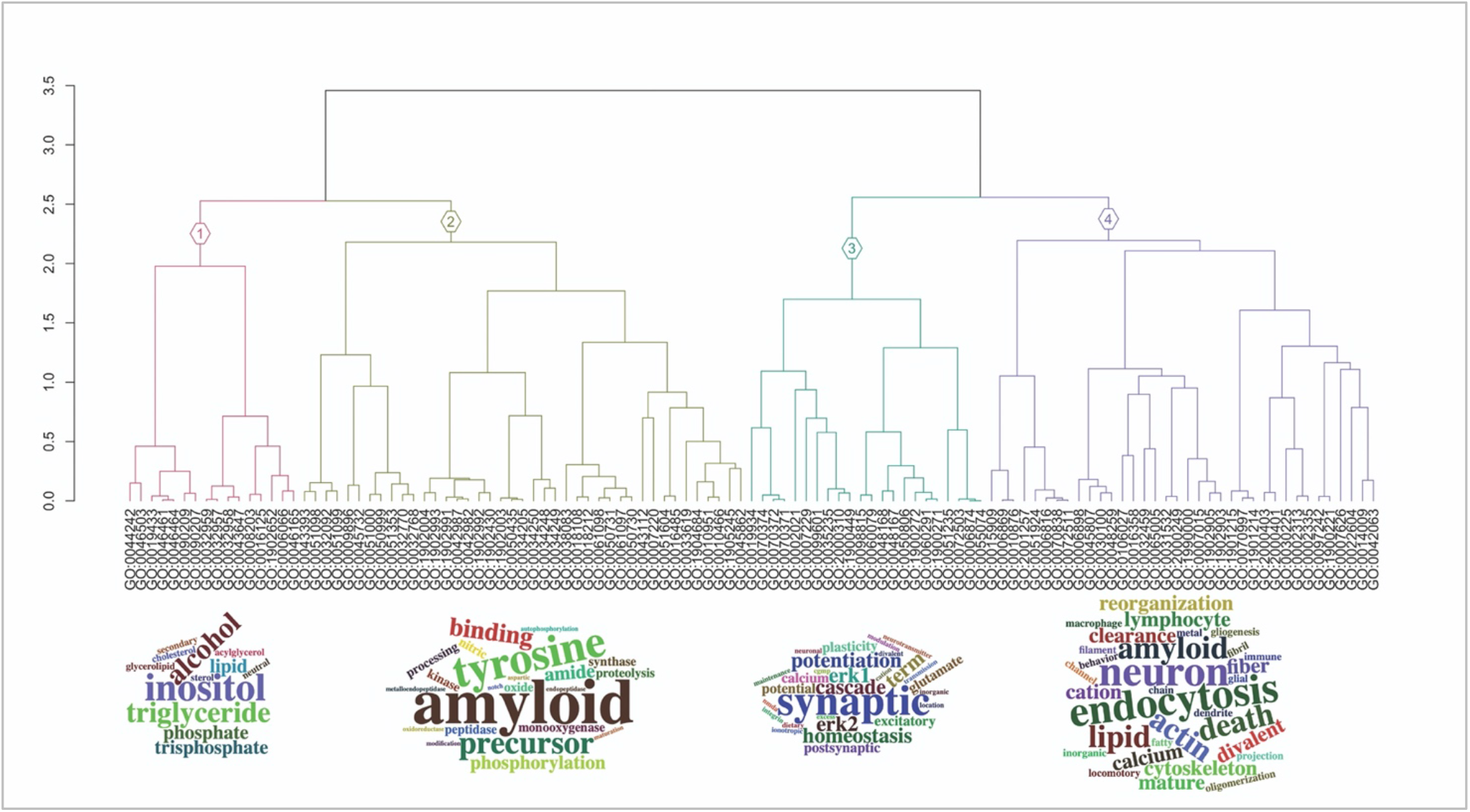
Hierarchical clustering of the semantic similarity matrix and the 4 functional clusters’ definition. Dendrogram of the hierarchical clustering analysis and the 4 functional clusters, along with word-clouds of the most frequent terms per cluster. Hierarchical clustering was performed on the semantic similarity distance matrix (using *Lin* as semantic similarity metric). We used the dynamic tree-cut method to define the number of functional clusters, specifying 15 as the minimum number of terms per cluster. We then used word-cloud visualization as well as manual interpretation of the biological processes underlying each functional cluster to label each cluster to *Lipid/Cholesterol metabolism* (cluster 1), *β -Amyloid metabolism* (cluster 2), *Synaptic plasticity* (cluster 3) and *Endocytosis/Immune signaling* (cluster 4).

## Supplementary Methods

### Populations

The 100-plus Study focuses on the biomolecular aspect of preserved cognitive health until extremely old ages. This study includes (1) Dutch-speaking centenarians who can (2) provide official evidence for being aged 100 years or older, (3) self-report to be cognitively healthy, which is confirmed by an informant (*i*.*e*. a child or close relation), (4) consent to donation of a blood sample and (5) consent to (at least) two home-visits from a researcher, which includes an interview and neuropsychological testing.^14^ This study also includes (1) siblings or children from centenarians who participate in the 100-plus Study, or partners thereof who (2) agree to donate a blood sample, (3) agree to fill in a family history, lifestyle history, and disease history questionnaire. The Longitudinal Aging Study of Amsterdam (LASA) is an ongoing longitudinal study of older adults initiated in 1991, with the main objective to determine predictors and consequences of aging.^19,20^ The SCIENCe is a prospective cohort study of subjective cognitive decline (SCD) patients.^21,22^ Participants undergo extensive assessment, including cerebrospinal fluid collection (CSF) and optional amyloid positron emission tomography scan (PET), with annual follow-up. The primary outcome measure is clinical progression. All individuals were labeled cognitively intact. The Netherlands Brain Bank (NBB) cohort is a prospective donor program for psychiatric diseases. All subjects were labeled cognitively intact after neuropathological examination.^23^ The Netherland Twin Registry study (NTR) was established in 2004 to collect biological and environmental data in twin families to create a resource for genetic studies on health, lifestyle, and personality.^24^

### Genotyping and imputation

Genetic variants in our populations were determined by standard genotyping and imputation methods, and we applied established quality control methods: we genotyped all individuals with the Illumina Global Screening Array (GSAsharedCUSTOM_20018389_A2) and excluded individuals with low-quality genotypes (individual call rate <98%, variant call rate <98%), individuals with sex mismatches and variants deviating from Hardy-Weinberg equilibrium (*p*<1×10^−6^). Genotypes were prepared for imputation comparing variants identifiers, strand and allele frequencies to the Haplotype Reference Panel (HRC v1.1, April 2016), and all remaining variants were submitted to the Sanger imputation server (https://imputation.sanger.ac.uk).^53^ The server uses EAGLE2 (v2.0.5) to phase the data, and imputation to the reference panel was performed with PBWT.^54,55^ Before analysis, we excluded individuals of non-European ancestry and individuals with a family relation, leaving 2,905 population subjects and 343 cognitively healthy centenarians for the analysis.

### Variant-gene mapping

We annotated each variant to the likely affected gene(s), so-called *variant-gene mapping*, combining annotation from *Combined Annotation Dependent Depletion* (CADD, v1.3), *expression-quantitative-trait-loci* in the blood (eQTL from GTEx v8), and positional mapping (from RefSeq build 98).^56–58^ In the case of coding variants, we confidently associated the variant with the corresponding gene. Alternatively, we first considered possible eQTL associations. When these were not available, we included all genes at increasing distance *d* from the variant (starting with *d* ≤ 50*kb*, up to *d* ≤ 500*kb*, increasing by 50*kb* until at least 1 gene was found). Our procedure allows the association of each variant with one or multiple genes (*Figure S2*).

### Gene-pathway mapping

The resulting list of genes was used to find the molecular pathways enriched in the AD variants. See *Figure S2* for a schematic representation of our annotation framework. We realized that allowing multiple genes to associate with each variant could result in an enrichment bias, as neighboring genes are often functionally related. To control this, we implemented a sampling technique: at each iteration, we (*i)* sampled one gene from the pool of genes associated with each variant, and (*ii*) performed a gene-set enrichment analysis with the resulting list of genes. The gene-set enrichment analysis was performed considering biological processes (BP) and implemented with the *enrichGO* function of the R package *clusterProfiler*, with all genes as background and correcting p-values controlling the False Discovery Rate (FDR). Finally, we averaged p-values for each enriched term over the iterations (N=1,000). To facilitate interpretation, we merged significantly enriched biological processes. First, we calculated the semantic similarity between all significant biological processes (*i*.*e*. FDR<5%) using *Lin* as a distance measure.^59^ We then applied hierarchical clustering on the resulting distance matrix and selected the number of functional clusters using the dynamic tree-cut method as implemented in *cutreeDynamic* function from the R package *WGCNA*, specifying 15 as the minimum number of terms per cluster (using the default value of 20 resulted in 2 functional clusters only). To provide an interpretation of each functional cluster, we selected the most frequent words describing the biological processes underlying each cluster, and show this as word-clouds as implemented in R package *wordcloud2*. Finally, by counting how often a functional cluster was associated with a gene, we could calculate a weighted annotation of each gene to the 4 functional clusters, so-called gene-pathway mapping (*Figure S2*).

Due to the initial selection of significantly enriched BP, not every gene in the list of variant-associated genes is annotated with (at least one of) these terms. Consequently, these genes could not be related to the final functional clusters. To overcome this, we connect these genes to the functional clusters using a k-nearest neighbor (k-NN) imputation. The k-NN model was initially trained using the functional clusters as classes and the semantic similarity matrix between the enriched biological processes as features (*feature terms*). Then, for each gene with missing annotation, we (*i*) extracted all the biological processes the gene is involved in (*input biological processes*), and (*ii*) calculated the semantic similarity matrix between these terms and the *feature terms*, which defines the similarity between the *input biological processes* and the *feature terms*. Finally, we (*iii*) predicted the probability of classification of the similarity matrix to the classes (functional clusters), and used this as weight for the gene-pathway mapping (*Figure S2*).

### Variant-pathway mapping

The variant-pathway mapping represents the combined annotation of each variant to the different functional clusters. As such, it depends on the variant-gene mapping and the gene-pathways mapping. Briefly, given a variant *k*, we (*i*) retrieved all the genes that were associated with the variant in the variant-gene mapping, *G*_*k*_, and (*ii*) retrieved all the biological processes (gene ontology term identifiers) that were associated with these genes, *GO*_*G*_. Because we clustered biological processes into functional clusters, by looking at which functional clusters the *GO*_*G*_ belonged to, we could assign a weight of association for variant *k* to each of the functional clusters.

### Variant-cell-type mapping

To study brain-specific cell-types and their relationship with AD-associated variants, we used the publicly available gene expression dataset GSE73721: this dataset includes gene expression values of 6 fetal astrocyte samples, 12 adult astrocyte samples, 8 sclerotic hippocampal samples, 4 whole human cortex samples, 4 adult mouse astrocyte samples, and 11 human samples of other purified central-nervous-system (CNS) cell-types. We restricted to the gene expression of 12 astrocyte samples and 11 samples of purified CNS cell-types from the cortex of adult humans (total N=23, mean age of 41.5±19.6 years). To calculate the variant-cell-type mapping, we averaged the gene expression of the genes mapping to the same variant.

## Notes

### Competing Interest Statement

The authors have declared no competing interest.

### Author Declarations

The Medical Ethics Committee of the Amsterdam UMC (METC) approved all studies. All participants and/or their legal representatives provided written informed consent for participation in clinical and genetic studies.

